# A Formal Model of Mood Disorders Based on the Neural Circuit Dynamics of the Triple Network Model

**DOI:** 10.1101/2024.06.16.24309000

**Authors:** Alan Lawrence Rubin, Mark Walth

**Affiliations:** 1233 Beech Street, Atlantic Beach, New York, 11509; Department of Mathematics, Cornell University

## Abstract

Psychiatric diagnoses are based on consensus and are not related to pathophysiology, leading to confusion in treatment and in basic and clinical psychiatric research. The pathology of mood disorders arises from the intrinsic function and interactions between key neural circuits of the triple network. These circuits are the central executive network composed of the dorsolateral prefrontal cortex and posterior parietal cortex; the default mode network consisting of the dorsal medial prefrontal cortex, posterior cingulate/precuneus and angular gyrus and the salience network made up of the anterior insula, dorsal anterior cingulate cortex associated with subcortical limbic nodes including the amygdala. In this work, we develop a formal model using nonlinear dynamics and network theory, which captures the dynamic interactions of these three brain networks, allowing us to illustrate how various mood disorders can arise. Recurrent circuit dynamics are modeled on the physio-dynamics of a single neural component and is dependent on a balance of total input (feedforward and feedback) and the sensitivity of activation of its neural components. We use the average percentage of maximal firing rate frequency as a measure of network activity over long periods, which corresponds to fMRI activity.

While the circuits function at moderate rates in euthymia, depressive symptoms are due to hypoactivity of the CEN and SN and hyperactivity of the DMN. Mania arises from a hyperactive SN with hypofunction of the CEN and moderate to high activity of the DMN. Functional abnormalities arise from genetic or epigenetic changes, affecting either the weight of neural interconnections or the sensitivity of activation of neurons comprising the network. Decreased excitation in unipolar depressive states is caused by diminished dendritic branches and decreased density of AMPA and NMDA receptors or a decrease in glutamate released by presynaptic neurons. All bipolar states result from heightened neural sensitivity due to altered sodium, calcium, or potassium channel conductance. Our formal model of mood disorders is consistent with fMRI studies, genetic research, as well as preclinical and clinical studies.

## Introduction

Current psychiatric diagnoses, including diagnoses of mood disorders, are based on criteria established by expert consensus rather than recognized pathophysiologies [1–2]. As a result, the same diagnosis can be used for conditions which differ in symptomatology, frequency, duration, and level of disability while different diagnoses may share similar symptomatology. Since medications often work as neuromodulators, abnormality of neuromodulation has been the prevailing hypothesis of psychopathology [3]. Researchers have increasingly questioned its validity [4–7].

This lack of understanding of pathophysiology leads to heterogeneity in defined patient populations, causing confusion in selecting subjects for research studies and in selecting effective treatments for individual patients and leading to a failure of innovative treatments [8,9]. As a result, a low percentage of patients diagnosed with major depression responded to their initial and subsequent treatments according to one often quoted multicenter study [10].

We are proposing a formal model of mood disorders based upon dynamical systems interaction as an alternative to the neuromodulator theory. A recent assessment of existing formal models of mood disorders by Nunes proposed that an effective dynamical model must satisfy three validity criteria: face, predictive and construct validity. A valid model should be able to identify the target symptoms, predict the onset and transitions between symptom states and provide a plausible mechanism for mood states and transitions [11]. We suggest that our model satisfies these three criteria.

Over the past two decades significant progress has been made in understanding the function and interactions of brain centers and in the application of neural network theory, matrix connectivity and information theory to brain function. These mathematical formulations offer a foundation for understanding the interactions of brain centers and their interconnections which can be used to identify the activity of neural circuits causing mood disorders [12–18]. Using network theory, we can conceptualize the vast complexity of the brain as an array of nodes connected by large bundles of myelinated tracts or edges. Nodes with complementary functions are connected into larger networks in a hierarchical manner with each later node refining the output of the prior. As will be explained in greater detail in the methods section, we conclude that the weight of evidence favors the hypothesis that mood disorder symptoms largely reflect the functioning of the central executive network (CEN), the salience network (SN), and the default mode network (DMN) [19–21].

The CEN, also called the frontoparietal network, is composed of the dorsolateral prefrontal cortex and the posterior parietal cortex. It is responsible for higher reasoning, problem solving, selective focusing, and choice of alternative actions based on an assessment of likely outcome. It also suppresses the immediate expression of more primitive impulses [22]. A loss of CEN leads to a loss of focus and reasoning and greater impulsivity [23,24].

The exact constituents of SN differ among researchers, but all agree that the primary hubs are the anterior insula and the anterior cingulate cortex which relate to the amygdala and hippocampus in association with the cortico-striato-thalamo-cortical circuit (CSTC) [25,26].

The SN is responsible for identifying an object of interest, assessing its value (attractive or aversive) and selecting an immediate response to salient stimuli. It may also through its interconnections be involved in self-awareness, communication, the integration of internal and external sensation, and may activate either the CEN or the DMN. The CEN is, therefore, dependent on the SN for its function [27–28]. Depression of SN-CEN circuit function leads to a muted CEN response to input which may include indifference [29]. Extreme SN activity leads to the characteristic symptoms of mania: hyperkinetic movements, euphoria, promiscuity, and reckless risk taking-based on a summary by Cotovio et.al. who reported hyperactivity of the ACC, amygdala, and basal ganglia which are components of SN-CSTC [30].

The default mode network (DMN) comprises the medial prefrontal cortex, posterior cingulate, precuneus and angular gyrus [32]. It is so named because it is active when the CEN and SN are inactive and is involved in rumination, introspection, self-reference, and daydreaming, while DMN hyperactivity causes depressive brooding as well as psychosis in depression, mania, and schizophrenia [33–35].

Genetic studies of bipolar disorder consistently identify alleles of ANK3, CACNA1C, and KCNB1 [36–42]. CACNA1C and KCNB1 are components of calcium and potassium channels, respectively. {39-42]. ANK3 controls the density and correct placement of sodium, potassium and calcium ion channels at the axon hillock, Nodes of Ranvier and apical dendrites [42–45,45,47]. Although these alleles are present in only a small percentage of subjects with bipolar disorder, they nevertheless suggest a pathophysiology that may result from small effects of multiple other alleles affecting ion conduction. Ion conduction underlies neural sensitivity to excitation and in bipolar disorder causes a high sensitivity to neural input [36,37,45–50].

There has been no allele consistently identified for depression [51]. However, human and animal studies have shown increased dendritic sprouting and increased dendritic volume in both depressed human subjects and animals treated with antidepressants, leading to the hypothesis that depression results from decreased postsynaptic density of AMPA or NMDA receptors [52–58].

To our knowledge this paper is a unique attempt to present a formal model of mood disorders based on the triple circuit model and grounded in neurophysiology and genetics. Starting with a simple model of single neuron dynamics, our model is consistent with fMRI studies of brain center activity and connectivity, the effects of gene variants on ion channels, as well as preclinical and clinical research.

## Method

In the method section, we identify and justify the underlying assumptions of the formal model by providing their grounding in neurophysiology, fMRI studies and in clinical observation. These assumptions are:

1. Mood disorders are disorders of neural circuits.
2. Based on current research the most likely circuits are the CEN, SN-CSTC (SN combined with nodes of the cortico-striatal thalamic circuit) and DMN.
3. The activity of each network is an average of network frequencies over prolonged periods (weeks or months).
4. Since these are recurrent circuits the average activity of the circuit can be modeled on the dynamics of a single neuron which is a component of the circuit.
5. The key variables of neural function are the strength of inputs, and the neuron’s sensitivity to inputs, which is determined by sodium, potassium, and calcium channel conductance.
6. Ion channel conductance is determined by genetics or epigenetics.
7. Mood states are not attributable to a specific node but arise from the combined interaction of individual nodes tied to their circuits and the interactions of those circuits.
8. The symptoms of mood disorders are caused by dysfunction within networks and between interacting networks.
9. Depressive episodes are due to hypofunction of the CEN and SN-CSTC and manic episodes are due to excessive activity of SN-CSTC and hypofunction of the CEN.
10. Bipolar disorder states are due to channelopathy.
11. The interactions of the networks are best quantified by using the nonlinear dynamics of neuron firing rates.

### Review of fMRI brain center activity in mood disorder states

FMRI records activity of specific brain centers which are the major nodes of neural networks, and coincidence of nodal activity is a marker for the connectivity of their neural networks.

Patterns of activity of key brain centers in depression and bipolar mood states were determined by reviewing fMRI studies of mood disorders by searching Pub Med and Google Scholar. For depression, we used only studies of currently depressed subjects. For bipolar disorder we used studies that clearly indicated current depression or mania and attempted to determine if the subjects were medicated. We also searched for studies of fMRI activity of the centers which are key to mood disorders: ventrolateral prefrontal cortex (VLPFC), dorsolateral prefrontal cortex (DLPC), medial prefrontal cortex (MPFC), anterior insula (AI), anterior cingulate cortex (ACC), amygdala, thalamus, hippocampus, and striatum, which will be reviewed below.

Systematic review of two decades of fMRI depression studies reveals inconsistent findings due to small sample size, heterogeneity of patient samples, medication, differences in protocol e.g., resting state versus task oriented, and technical difficulties such as noise to signal ratio. The preponderance of evidence supports a loss of function of the prefrontal cortex, as well as decreased activity or connectivity of the AI, ACC, ventral striatum, basal ganglia, and medial thalamus [59–67]. In depressed subjects the amygdala may be hyperactive in tasks which evoke negative emotion [68–69].

FMRI studies of mania are contradictory due to the impracticality of obtaining compliance from unmedicated acutely manic subjects. fMRI studies show hyperactivity of the amygdala, basal ganglia, and ACC based on a summary of studies using varied methodologies by Cotovio, as well as increased ACC and caudate activity in a small study by PET scan [53–54]. Studies of functional connectivity in mania, while generally agreeing with a decreased VLPFC, and increased amygdala activity, as well as decreased suppression of amygdala hyperactivity by the VLPFC, nevertheless, do not require actively symptomatic or unmedicated subjects, often do not specify bipolar state, use small numbers of subjects, and use highly divergent methodologies [70–74].

In identifying candidate networks, we used a simplifying assumption that in a recurrent network the dominant node, with greatest effective connectivity, should set the prevailing frequency over prolonged time periods. We found that the crucial nodes for mood disorders most frequently identified by fMRI studies were constituent nodes of the CEN, DMN and SN combined with the CSTC [59–76]. Also, connectivity studies confirmed the presence or absence of an inter-connection between the key nodes of the CEN, DMN and SN-CSTC (fig. 1) and supports the conclusion that abnormalities in the function of one circuit leads to abnormal function of any circuit with which it interacts [59–67, 74–79]. Therefore, the symptoms of mood disorders arise from the disordered function of one or more networks and their interactions. The fMRI studies indicate that depressive symptoms result from low activity of key nodes of the CEN and SN and increased activity of the DMN [77–79]. In the depressive state the amygdala may be hyperactive but has decreased functional connectivity with other nodes of the SN-CSTC [80, 81]. The manic state results from an unrestrained SN-CSTC with subdued CEN [30, 31].

**Fig 1.**
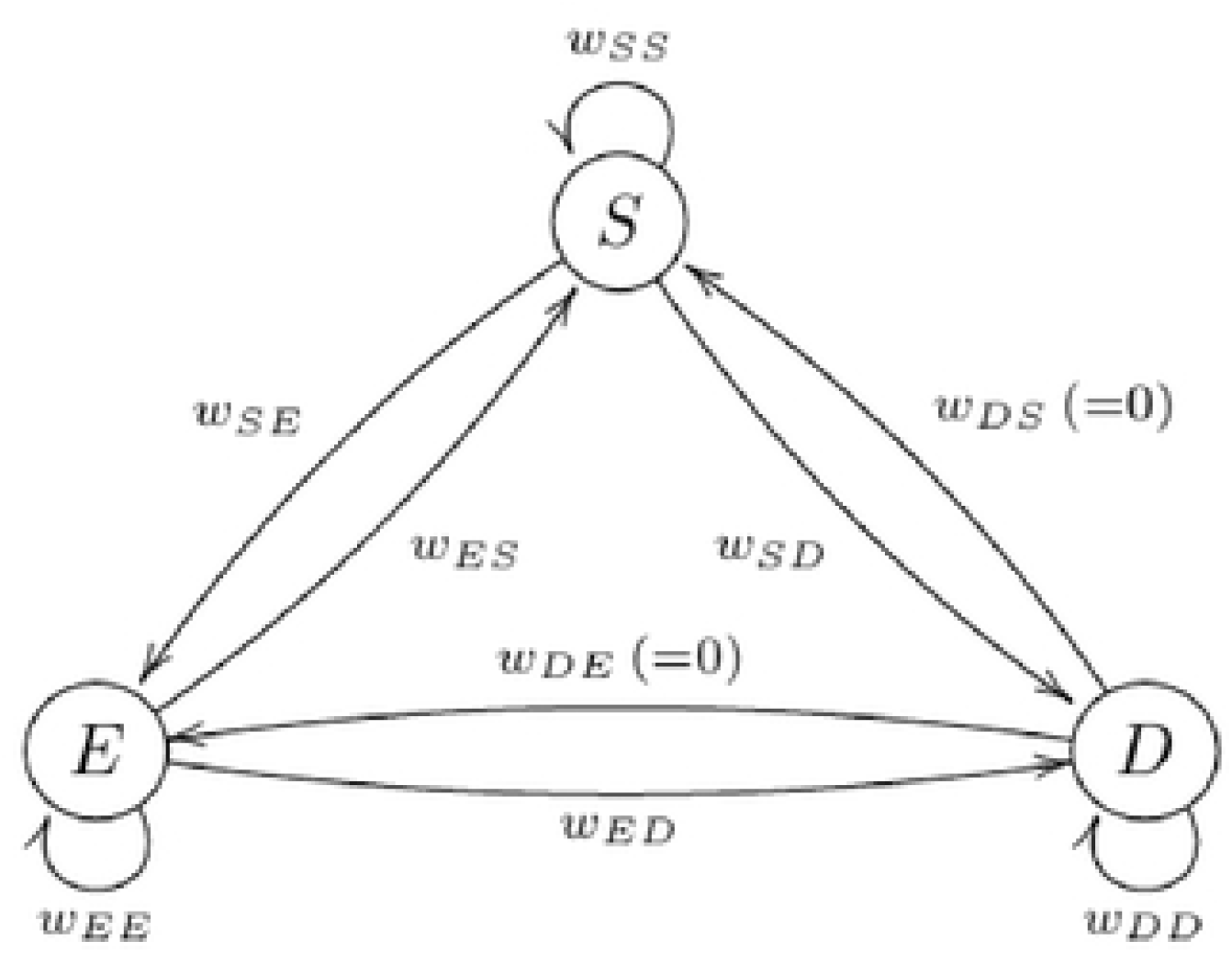
Diagram of triple circuit dynamics. A schematic model showing the interconnections of the three network model.

In our model of bipolar disorder, we assume a channelopathy affecting SN which causes an excessive sensitivity to change in SN recurrent activity [43–50].

### Network Model

Differential equations can be used to model the dynamics of the networks with greater clarity and predictive power because recurrent and internetwork output evolve over time. The network is modeled as a weighted directed graph, with weightings representing the strength of the connections within and between the individual nodes in the network. These weightings appear in the governing differential equations in the form of the connectivity matrix [13].

Network activity is specified as a relative firing rate. How information is encoded and transmitted by neurons is under active investigation [82]. However, for our purposes we are more interested in whole circuit activity as measured by the metabolic needs of the constituent nodes and dependent on the percentage of activated neurons and their firing rates, as captured by fMRI. This activity would be averaged over periods of weeks or months, typical of episodes of mood disorders.

We use an equation for single neuron dynamics [35,36] to represent the activity of the entire circuit, simplifying circuit dynamics by making the network frequency dependent on only two parameters- the average neuron’s weighting (*W*), given by the adjacency matrix, and sensitivity to stimulation (σ) [35,36] (fig. 1). In our simple model, all networks have the same σ for euthymia and unipolar depression, however, for all bipolar states SN has a greater σ.

Positive and negative weightings indicate excitatory or inhibitory connections. We use the convention that *W*ij denotes the influence that network i has on network j. For example, *W*SE denotes the influence of SN on CEN. The assigned weighting of intra-network stimulation (e.g., *W*SE) and inter-network stimulation (e.g., *W*ss) is the sum of all excitatory and inhibitory weightings of feedforward and feedback connections.

We use weightings to produce levels of network activation that model the fMRI results of network nodes associated with the specified mood disorder. In our model all network variables must be relative since the actual values for average frequency, weightings, and slope of activation in mood states are not known.

For all unipolar mood states all networks are at homeostatic equilibrium: If their firing rates are altered, they will shortly return to their fixed firing rate. However, for all bipolar states we assign less stable fixed points to produce the rapid and dramatic fluctuations in mood characteristic of bipolar disorder. To create relatively unstable fixed points, we increase the σ of SN. This implies higher sodium or calcium conductance or lower potassium conductance in all bipolar states.

The matrix *W* gives the weightings of inter and recurrent stimulation of the networks and follows the conclusions regarding the interactions of the triple circuits. We assigned a relatively lower recurrent weighting to the CEN, making the CEN more vulnerable to changes in inter-network excitation. Other rules for network interconnections are: SN is the primary driver, it can increase or decrease SN-CEN and SN-DMN; CEN weightings are only inhibitory; it can decrease SN and decrease DMN; DMN is passive, it is acted upon by SN or CEN. SN, through its dominant AI node, can turn on or off the CEN and DMN even when SN activity is high [27,28].

### Formulation of network equations

In the model, the strength of influence of unit *i* on unit *j* will be represented by a scalar called the *weighting,* which will be denoted *w*_*ij*_. For example, the strength of influence of the salience network on the central executive network is denoted *w*_*SE*_, and is represented in Figure 1 as the arrow from *S* to *E*. The governing equation of such a firing rate network is given by equation 7.10 of [12]:

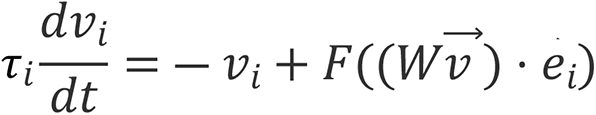

where:

- *v*_*i*_ represents the firing rate of unit *i*. For example, *v*_*S*_ denotes the firing rate of the salience network;
- *v* is a vector containing the firing rate of each unit, *v* = (*v*_*S*_,*v*_*E*_,*v*_*D*_);
- 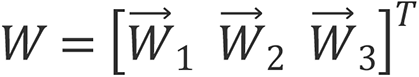 is the matrix containing the weightings of all of the connections within the network, and 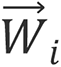 denotes the weightings of all connections going in to node *i*;
- *τ*_*i*_ is a time constant;
- *e*_*i*_ is the *i*^*th*^ standard basis vector;
- *F* is the “activation function” of the neurons, representing how the firing rate of a neural unit changes as a function of its input current.

We use the sigmoidal activation function

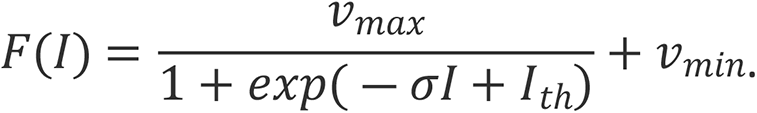

In this expression, *I* is the total input current to the neural unit, *v*_*max*_ is the maximum firing rate of the neural unit, *v*_*min*_ the minimum possible firing rate - which we take to be small but nonzero - *I*_*th*_ is the activation threshold, and *σ* is a parameter which controls the steepness of the activation curve. Biologically, the parameter *σ* represents the overall sensitivity of a neural unit to input current. In a biological neuron, sensitivity is determined by a range of factors, including ion channel density and synaptic sensitivity - in our model, *σ* represents an amalgamation of all such factors. A larger value of *σ* corresponds to a more sensitive neural unit, and vice versa. In some texts, see for example [12], the constants are taken to be *v*_*max*_ = 100Hz, *I*_*th*_ = 50nA, *σ* = 17.5, and *τ* = 5ms. We choose to scale our parameters in such a way that emphasizes qualitative behavior of the network, rather than focusing on specific quantitative relationships.

In this scaling, *v*_*i*_ represents the firing rate of unit *i*, represented as a fraction of its maximum possible firing rate. For example, if *v*_*i*_ = 0.6, this means that unit i is firing at 60% of its maximum possible firing rate. Finally, for simplicity, we take all the time constants to be equal and set them to *τ*_1_ = *τ*_2_ = *τ*_3_ = 5. Therefore, the parameters that we manipulate in this model are the weightings between neurons, *W*, and the activation sensitivity *σ*.

## Results

For the euthymic state, σ is set at 2.2 which represents moderate slope of activation to depolarizing current based upon the density of sodium, calcium and potassium channels at the axon hillock and axon terminals of all three networks (CEN, SN, DMN). In the euthymic state recurrent CEN weighting (*W*_EE_, 0.9) is far smaller than recurrent SN weighting (*W*ss, 2.0) causing CEN function to be highly dependent on SN input from AI. The euthymic state is homeostatic with stable intra and inter network activity of the three networks which all function at midrange activity. The connectivity matrix used in the euthymic state is given by: *W*ss 2.0, *W*_SE_ 0.8, *W*_SD_ 0.6, *W*_ES_ −3.5, *W*_EE_ 0.9, *W*_ED_ −0.5, *W*_DS_ 0, *W*_DE_ 0, *W*DD 1.5. The resulting behavior of the three networks is shown in Figure 2.

**Fig 2.**
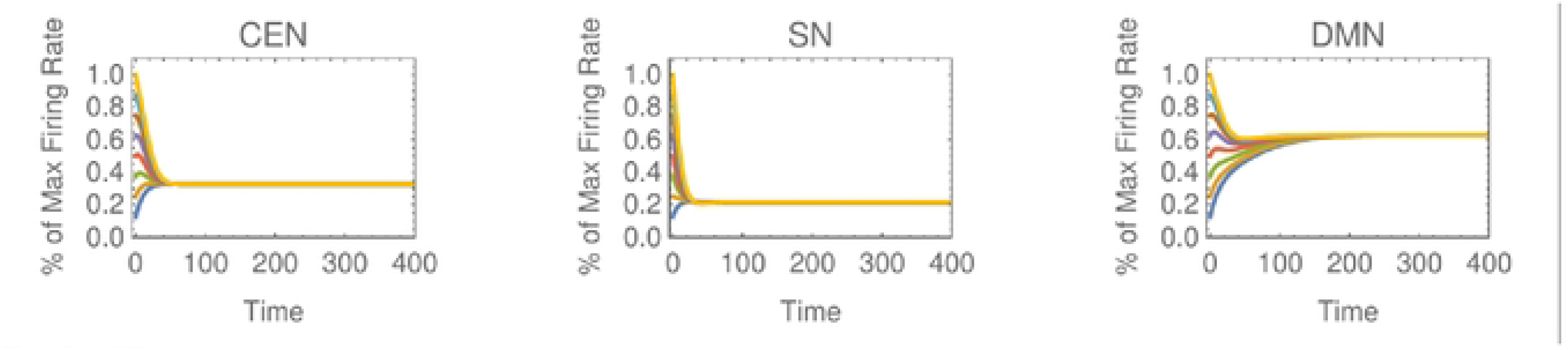
Euthymic. In the euthymic state, all three networks tend towards a stable fixed point at an intermediate firing rate, corresponding to a healthy homeostatic state.

For unipolar depression, σ remains at 2.2. The unipolar depressive syndrome is caused by a decrease in the SN recurrent stimulation by 25%. Although CEN network recurrent weighting remains constant, its activity is greatly reduced by decreased SN input. Decreased CEN activity leads to heightened DMN activity of 13%. The connectivity matrix for this state is given by *W*_SS_ 1.5, *W*_SE_ 0.8, *W*_SD_ 0.6, *W*_ES_ −3.5, *W*_EE_ 0.9, *W*_ED_ −0.5, *W*_DS_ 0, *W*_DE_ 0, *W*_DD_ 1.7. The network dynamics of the unipolar state are shown in Figure 3.

**Fig 3.**
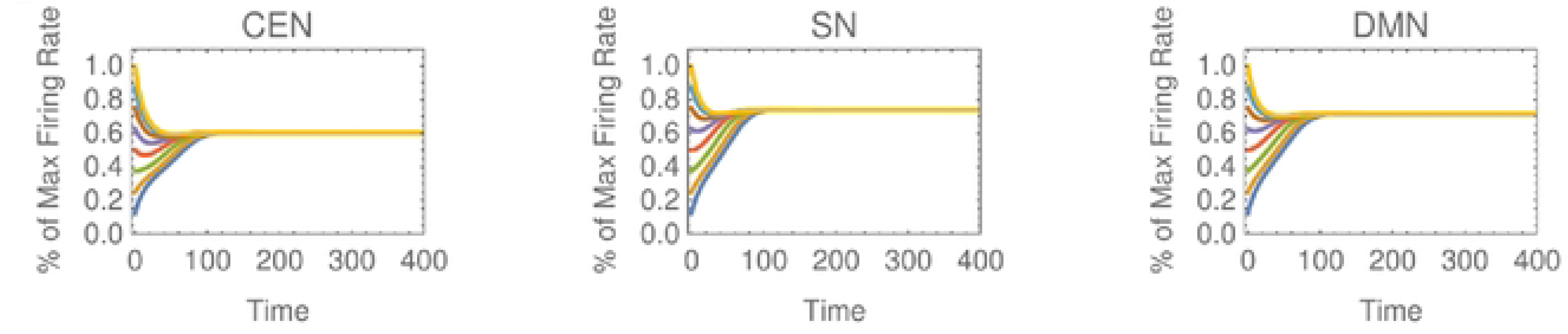
Unipolar depressed. In unipolar depression, the salience network and the central executive network are both operating at an extremely low activity level, while the default mode network maintains an elevated level of activity.

For bipolar disorder, the SN σ is increased to 2.5 in all bipolar states, 13.6% higher than the SN σ for euthymia and unipolar depression, while CEN σ and DMN σ remain at 2.2. (fig. 4-5) This small increase bipolar SN σ causes extreme responses in SN activation due to small changes in SN recurrent excitation. The bipolar euthymic state requires a 71% increase in inhibition of CEN-SN and decreased SN-CEN excitation of 25%. The connectivity matrix used to generate this state is: *W*_SS_ 2, *W*_SE_ 0.6, *W*_SD_ 0.6, *W*_ES_ −0.6, *W*_EE_ 0.9, *W*_ED_ −0.5, *W*_DS_ 0, *W*_DE_ 0, *W*_DD_ 1.5. The network dynamics are illustrated in Figure 4.

**Fig 4.**
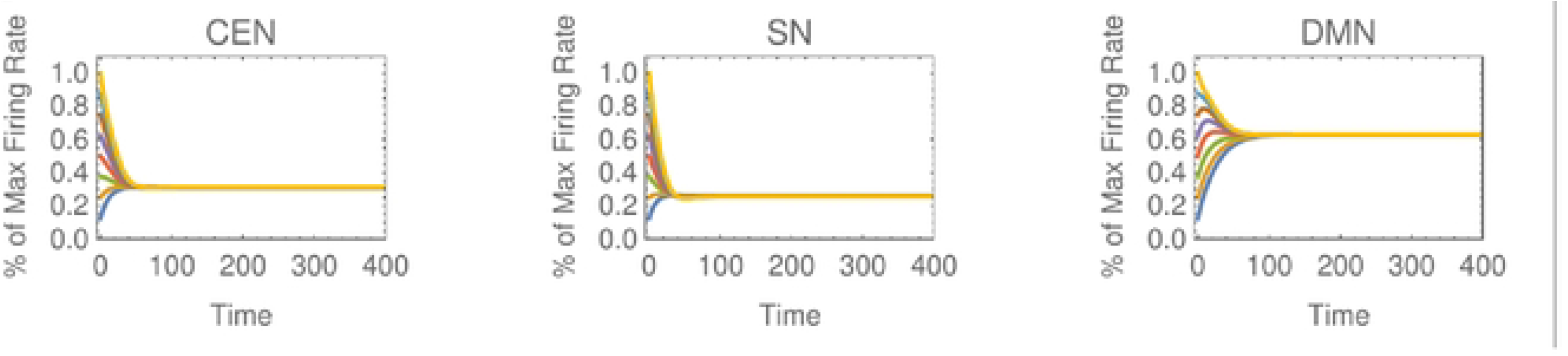
Bipolar euthymic. The euthymic bipolar state is indistinguishable from true euthymic, with similar activation of each of the three networks. Bipolar depression is caused by a minor change in the network weightings: The activity of the salience and executive networks collapse, while the default mode network maintains high activity. The sensitivity to network parameters is caused by the increased sensitivity σ of the salience network.

Bipolar depression results from a 10% reduction of SN recurrent excitation. The connectivity matrix is: *W*_SS_1.8, *W*_SE_ o.6, *W*_SD_ 0.6, *W*_ES_ −0.6, *W*_EE_ 0.9, *W*_ED_ −0.2, *W*_DS_ 0, *W*_DE_ 0, *W*_DD_ 1.5. The network dynamics are shown in figure 5.

**Fig. 5.**
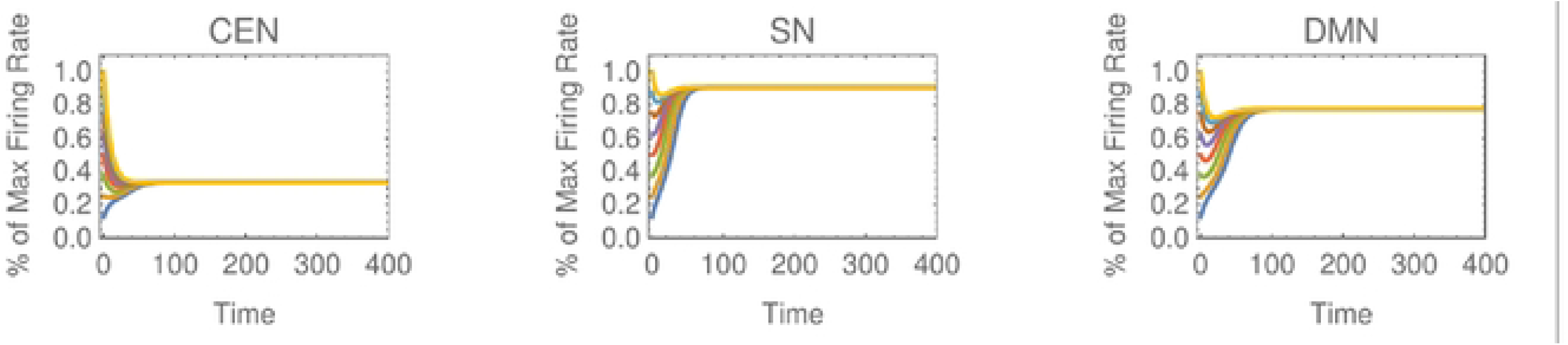
Bipolar Depressed. Bipolar depression is caused by a minor change in the network weightings: The activity of the salience and executive networks collapse, while the default mode network maintains high activity. The sensitivity to network parameters is caused by the increased sensitivity σ of the salience network.

Bipolar mania is caused by reduction of SN-CEN excitation by 50% and a corresponding increased SN-DMN excitation of 33%. The connectivity matrix is: *W*_SS_ 2.0, *W*_SE_ 0.2, *W*_SD_ 0.4, *W*_ES_ −0.6, *W*_EE_ 0.9, *W*_ED_ −0.5, *W*_DS_ 0, *W*_DE_ 0, *W*_DD_ 1.5. The network dynamics are shown in figure 6.

**Fig 6.**
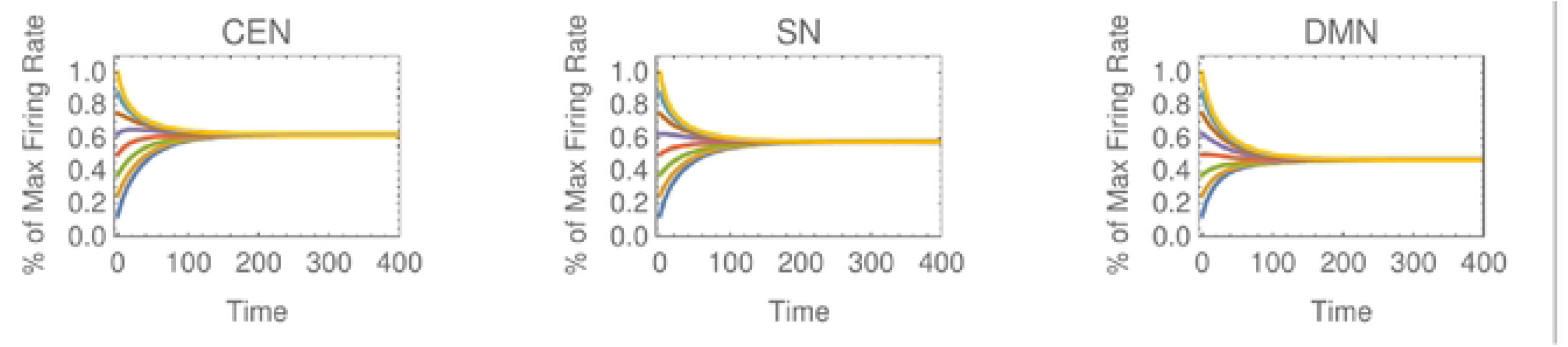
Bipolar manic. A slight change of the network weightings causes the bipolar-euthymic state to enter a manic state, in which the executive network is depressed, and the salience and default mode networks have extremely heightened activity.

Therefore, our model attributes both unipolar and bipolar depression to an initial loss of recurrent excitation of the SN and mania to a shift in SN excitation from CEN to DMN. Although not illustrated, excessive DMN activity may arise from increased *W*_DD_, increased *W*_SD_ or decreased *W*_ED_ (through decreased CEN-DMN inhibition) and produces psychotic symptoms [34,35].

## Discussion

We have presented what we believe is a unique formal model of mood disorders arising from the dynamics of coupled neural circuits that serves as an alternative paradigm to neuromodulation theory. It grounds mood states in a model which is consistent with what is known of neurophysiology and genetics and its assumptions are clear and capable of validation. It presents mood disorders as a group of symptoms arising from fluctuations of system parameters, much as the weather. While other formal models of the dynamics of mood disorders have been proposed, ours is unique because of its strong grounding in pathophysiology [11].

We demonstrate how intra-network recurrent excitation or inhibition can be based on the dynamics of neural circuits by using nonlinear dynamics and show how the resulting intra-network interactions of the CEN, SN-CSTC, and DMN networks lead to the pathologic circuit activity which underlies the symptom clusters of mood disorders.

The extreme complexity of brain function has presented a Gordian Knot which has been impossible to resolve. We have presented a radical simplification, comprising three circuits, their weightings of intrasynaptic strength and their neural sensitivity based on ion channel function. We model intra-circuit dynamics on recurrent networks: The entire network shares the dynamics of a single constituent neuron with all elements of a circuit conforming to the dynamics of a single nodal component. By simplifying an enormously complex problem we may have constructed a useful heuristic model for further research.

Any simplification opens itself to the criticism that it is overly simplistic and ignores important exceptions. In its defense we point out the extremely contradictory conclusions of decades of studies based upon small samples, different inclusion and exclusion criteria for subjects, the marked ambiguity of criteria for diagnoses, and the vagaries and artifacts of the emerging technologies of fMRI and genetic testing. With all the noise it is difficult to isolate a signal.

In constructing our model, we attempted to find the most replicated of conflicting results or a consensus opinion. Ultimately a researcher reviewing the studies must employ Bayesian probability as a tool of analysis, factoring one’s prior assumptions when weighing the likelihood of given test results- a technique which all seasoned clinicians employ in their daily practice.

Ultimately the question is, does this model work? Are its underlying assumptions provable? Does it lead to research that discovers previously unknown truths? Does it lead to new treatments? Does it lead to better ways of constructing studies?

We propose that the underlying cause of both unipolar depression and bipolar disorder lies within SN and that the underlying pathophysiology of bipolar disorder is a heightened sensitivity of the SN resulting from abnormal conductance of sodium, calcium and/or potassium channels due to genetic or epigenetic influences [35–50]. Applying the neural circuit model, both the rapidity and frequency of mood changes in the bipolar states result from unstable fixed points created by the heightened sensitivity of the SN.

Our formal model explains the mechanism of action of most bipolar medications-lithium, sodium valproate and atypicals - as sodium channel blockers [83–87]. It predicts that channelopathies and an abnormally high σ will be found in the key nodes of the SN-amygdala in cadaver studies of Bipolar I subjects. In our model excessive excitation of the DMN results in psychotic symptoms and explains the shared genetics of schizophrenia and bipolar disorder to alleles which cause excessive excitation of the DMN [34–35].

Transcranial magnetic stimulation (TMS) may ultimately prove to offer a more specific treatment for mood disorders. TMS treatment of depression induces a current between the dorsolateral prefrontal cortex and regions of SN [88–89]. TMS with deep cranial penetration may be more effective by reaching the deeper brain structures of the SN [90–92].

An implication of the theory is the importance of selecting subjects with homologous phenotypes for clinical treatment studies. The original Feigner criteria emphasized the episodic nature of major depression and bipolar disorder have been considerably broadened allowing for marked heterogeneity among research subjects ([1,2,93,94]. Homogeneity in research studies is better insured by similar symptomatology in unmedicated subjects rather than similar but amorphous diagnoses. Treatment studies should also use subjects homologous for symptoms as well as chronicity and general level of function. For example, in the STAR D study there was a considerably better response to antidepressants in the subset of subjects with a work history [95].

FMRI is currently the most effective means of identifying network pathology. However, since abnormalities of circuit activation are state dependent and state specific, FMRI studies of all bipolar subjects, regardless of state, are misleading as are studies of depressed patients irrespective of symptom profile and medication usage. Ideally, fMRI studies would compare imaging during and after episodes in large groups of symptomatically similar subjects to identify state driven abnormalities. Treatment studies of mood disorders should use unmedicated subjects with similar pretreatment fMRI and standardized fMRI protocols.

The dynamic model explains the differing response of unipolar and bipolar depressives to the increased feedforward excitation caused by antidepressants: The increased SN sensitivity of bipolar patients to antidepressant causes a manic response.

In conclusion our formal model fulfills the requirements given by Nunes et.al, outlining the necessary elements of a useful model of mood disorders: face, predictive and construct validity [11]. Its assumptions are testable, and its conclusions may lead to improvements in diagnosis, and treatment and suggest novel directions for future research.

## Author Contributions

Alan Lawrence Rubin performed the research, identified the neural circuit model and the mathematical model used, and wrote the paper.

Mark Walth completed the mathematical analysis and designed the graphs.

## Data Availability

All data are available within the manuscript.

## Acknowledgments

None

